# A Scalable Framework to Integrate Social Determinants of Health into Disease Risk Models using Biobank Survey Data

**DOI:** 10.1101/2025.09.29.25336903

**Authors:** Jane Brown, Abhijith Biji, Kathleen Ferar, Vikas Pejaver, Eimear E. Kenny, Bian Liu, Samira Asgari

## Abstract

Complex diseases are a major global health burden, yet our ability to predict who is at risk remains limited. Risk is shaped by both heritable factors and non-genetic environmental, behavioral, and social determinants of health, but these are rarely modeled together. The growing availability of large-scale, multimodal biobanks creates new opportunities to integrate diverse data types into more accurate disease risk models. Here, we apply Multiple Correspondence Analysis (MCA) to over 100 environmental, behavioral, and social variables from the All of Us biobank (N = 171,614) to generate low-dimensional embeddings that quantify non-genetic risk for six common chronic conditions. These embeddings recovered known and novel risk factors and consistently improved prediction beyond demographics and polygenic scores (PGS), with contributions to model performance (ROC-AUC) ranging from 0.03 to 0.05. For five of six diseases, the gains from MCA embeddings surpassed those attributable to PGS. Genetic and non-genetic risks combined largely additively: we observed little evidence of interaction effects (ΔAUC < 0.001) and highly stable variant effect sizes when embeddings were included in genetic association models (*r* > 0.98). In summary, we introduce a scalable, interpretable framework that summarizes survey-based environmental, behavioral, and social factors without prior assumptions about disease-specific variables. Our results demonstrate that these non-genetic contexts substantially improve prediction while acting largely additively with genetic risk, clarifying the role of gene-environment interplay in complex disease and supporting more equitable and robust risk modeling across diverse populations.

## Introduction

Complex diseases are the leading cause of global morbidity and mortality, accounting for the majority of Disability-Adjusted Life Years worldwide, with projections indicating they will dominate the global burden of disease by 2050 ^1^. Early identification of at-risk individuals and timely intervention are essential for reducing this burden, improving population health, and containing healthcare costs. However, developing risk prediction strategies that capture the multifaceted nature of chronic disease remains a fundamental challenge ^2^.

The origins of complex disease risk lie in the intricate interplay between genetic predisposition and environmental, behavioral, and social determinants of health. Yet current prediction strategies typically examine these contributors in isolation. Genetic risk is often modeled through polygenic scores (PGS), while epidemiological models focus separately on environmental, lifestyle, and social exposures ^2,3^. This disconnect stems from two key limitations: the scarcity of data resources that comprehensively measure both genetic and non-genetic factors, and the lack of analytic frameworks capable of integrating these distinct data modalities into unified models of disease. The challenge is further complicated by the nature of non-genetic exposures themselves, which are numerous, highly correlated, and often measured inconsistently across different cohorts, making their combined influence difficult to quantify.

Nonetheless, jointly modeling genetic and epidemiological risk factors is critical for accurate disease prediction and effective early intervention. The emergence of large, diverse biobanks that systematically collect genomic, electronic health record (EHR), and survey data makes it possible to achieve this goal, with the potential to improve risk prediction, clarify how risk factors interact to shape outcomes, and uncover new insights into disease pathogenesis.

To this end, here we leveraged the unique combination of genetic and survey data available in the All of Us biobank to address this gap. We applied Multiple Correspondence Analysis (MCA) to more than 100 environmental, behavioral, and social determinants of health variables, generating low-dimensional, interpretable embeddings that capture patterns of non-genetic risk. These embeddings allowed us to quantify how non-genetic factors contribute to disease risk and examine their impact on modifying genetic risk for six common chronic diseases with well-established polygenic architecture and documented environmental influences, namely asthma, prostate cancer, breast cancer, chronic kidney disease, coronary heart disease, and hypercholesterolemia.

## Results

### MCA reveals structured patterns in behavioral and social determinants of health

We used the All of Us biobank (version 7) to obtain EHRs, genetic data, and comprehensive survey responses from 413,457 total participants. Among them, 171,614 had both EHR data and genetic data available (**Methods**). We assigned case status using previously described ICD codes for each disease ^4^ (**Methods, Table 1**). Individuals without the relevant ICD codes served as controls (**Table 1**).

**Table 1:** Cohort demographics. Numbers in each cell represent the total number of individuals matching the criteria in the post-QC cohort (e.g., Self-reported non-Hispanic Blacks who had asthma), numbers in parentheses indicate percentages for the corresponding total count. EUR: European genetic ancestry, AFR: African/American genetic ancestry, AMR: Hispanic/Latino genetic ancestry, OTH: Other genetic ancestries.

|  | Group | Overall | Asthma | Breast cancer | Prostate cancer | Chronic Kidney disease | Coronary heart disease | Hypercholesterolemia |
| --- | --- | --- | --- | --- | --- | --- | --- | --- |
|  | All | 171614 (100.0%) | 30996 (100.0%) | 6871 (100.0%) | 4425 (100.0%) | 16840 (100.0%) | 25573 (100.0%) | 43879 (100.0%) |
| Genetic ancestry | EUR | 101980 (59.4%) | 18059 (58.3%) | 5062 (73.7%) | 3445 (77.9%) | 10105 (60.0%) | 17524 (68.5%) | 31043 (70.7%) |
|  | AFR | 34507 (20.1%) | 7247 (23.4%) | 876 (12.7%) | 628 (14.2%) | 4156 (24.7%) | 4658 (18.2%) | 6328 (14.4%) |
|  | AMR | 29004 (16.9%) | 5000 (16.1%) | 737 (10.7%) | 286 (6.5%) | 2246 (13.3%) | 2855 (11.2%) | 5291 (12.1%) |
|  | OTH | 6123 (3.6%) | 690 (2.2%) | 196 (2.9%) | 66 (1.5%) | 333 (2.0%) | 536 (2.1%) | 1217 (2.8%) |
| Self-reported gender | Male | 64135 (37.4%) | 8386 (27.1%) | 108 (1.6%) | 4263 (96.3%) | 8511 (50.5%) | 13922 (54.4%) | 18375 (41.9%) |
|  | Female | 103143 (60.1%) | 21767 (70.2%) | 6599 (96.0%) | 52 (1.2%) | 7938 (47.1%) | 11064 (43.3%) | 24495 (55.8%) |
|  | Other | 4336 (2.5%) | 843 (2.7%) | 164 (2.4%) | 110 (2.5%) | 391 (2.3%) | 587 (2.3%) | 1009 (2.3%) |
| Age | < 40 | 37167 (21.7%) | 6257 (20.2%) | 131 (1.9%) | < 20 | 656 (3.9%) | 515 (2.0%) | 1528 (3.5%) |
|  | 40 to 65 | 73783 (43.0%) | 14386 (46.4%) | 2623 (38.2%) | 628 (14.2%) | 5763 (34.2%) | 8014 (31.3%) | 15713 (35.8%) |
|  | >= 65 | 60664 (35.3%) | 10353 (33.4%) | 4117 (59.9%) | 3790 (85.6%) | 10421 (61.9%) | 17044 (66.6%) | 26638 (60.7%) |
| Self-reported race/ethnicity | Non-Hispanic White | 96216 (56.1%) | 16972 (54.8%) | 4760 (69.3%) | 3226 (72.9%) | 9524 (56.6%) | 16467 (64.4%) | 29232 (66.6%) |
|  | Non-Hispanic Black | 29888 (17.4%) | 6132 (19.8%) | 749 (10.9%) | 555 (12.5%) | 3702 (22.0%) | 4086 (16.0%) | 5467 (12.5%) |
|  | Non-Hispanic Asian | 4481 (2.6%) | 452 (1.5%) | 157 (2.3%) | 39 (0.9%) | 223 (1.3%) | 362 (1.4%) | 876 (2.0%) |
|  | Hispanic or Latino | 30854 (18.0%) | 5452 (17.6%) | 821 (11.9%) | 351 (7.9%) | 2415 (14.3%) | 3102 (12.1%) | 5775 (13.2%) |

To capture non-genetic environmental, behavioral, and social determinants of health, we assembled data from four All of Us surveys (The Basics, Lifestyle, Healthcare Access and Utilization, and Social Determinants of Health surveys; **Table S1**) along with area-level information linked to participants’ three-digit ZIP codes. This yielded 192 candidate variables across our 171,614 participants. After quality control (QC), 142 variables remained, comprising 136 survey questions and six area-level variables (**Tables S1-3, Methods**). Participants answered an average of 63.2 questions, though with substantial variation (standard deviation (SD) = 42.3, **Table S1**).

Applying Multiple Correspondence Analysis (MCA) to the post-QC categorical variables revealed distinct patterns in the low-dimensional space. The first axis explained 10.5% of the variance, while axes 1–10 together explained 23.8%, and axes 11–20 contributed an additional 6.1% (**Fig. S1**). Beyond this point, the variance explained plateaued (**Fig. S1**); therefore, we retained the first 20 MCA axes for downstream analyses. As expected for survey data capturing related constructs, responses showed high correlation, particularly within individual surveys (**Fig. 1A**).

**Fig. 1.**
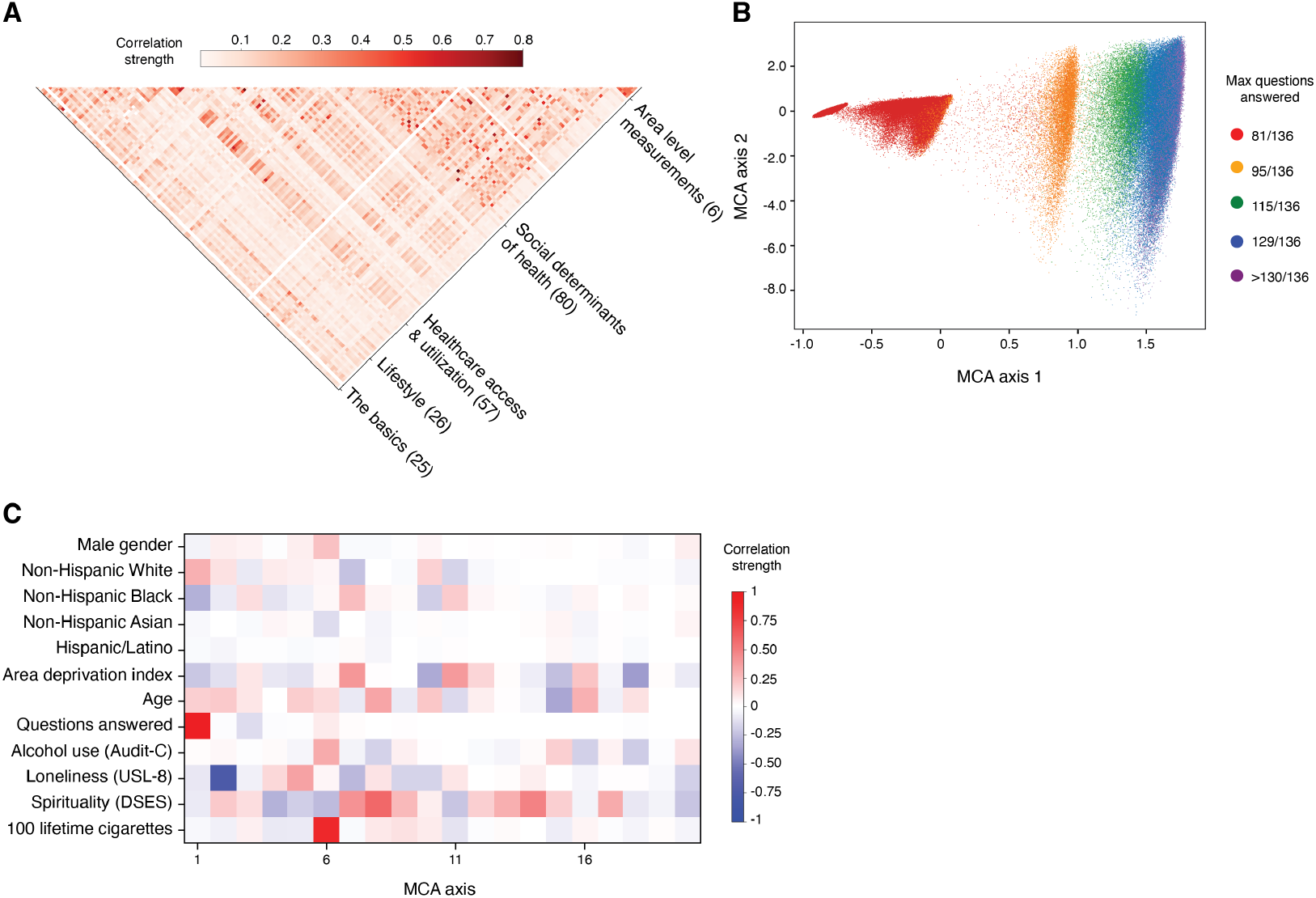
Survey variables and MCA-derived embeddings. **(A)** Pairwise correlations among post-QC survey questions used as input to MCA. Colors represent correlation strength (Cramer’s V), ranging from 0 (white) to 1 (dark red). Numbers in parentheses shows the number of post-QC question per survey. **(B)** Row projections of participants along MCA axes 1 and 2. Axis 1 is strongly associated with survey completion: individuals answering fewer questions cluster on the left, while those with more complete responses spread to the right. Each point represents one participant, colored by the maximum number of survey questions answered. **(B)** Correlations between the first 20 MCA axes and selected demographic, environmental, social, and behavioral variables. Colors reflect correlation magnitude, with Pearson’s correlation used for continuous variables (e.g., age, number of questions answered) and Point-Biserial correlation for binarized input variables.

MCA axis 1 did not appear to capture a single environmental, behavioral, or social determinant of health. Instead, it seemed to reflect participants’ broader socioeconomic and sociodemographic profiles (**Fig. S2**). Notably, this axis showed a very strong correlation with survey completion rates (Pearson *r* = 0.98, *p* < 1 × 10^−250^; **Fig. 1B**) and with demographic characteristics, including higher proportions of participants who self-identified as White, lower proportions who self-identified as Black, older age, and fewer who self-identified as male (*r* = 0.30, -0.18, 0.056, -0.057, respectively; all *p* < 1 × 10^−287^; **Fig. 1C**), even though neither survey completion nor demographic variables were used as inputs to the MCA. Taken together, these findings suggest that MCA axis 1 reflects underlying sociodemographic and response-completion structure, consistent with previous reports linking nonresponse to older age, lower socioeconomic status, and poorer health ^5–7^.

The remaining axes (2–20, cumulative variance explained = 19.4%) revealed patterns of co-occurrence between response categories and revealed distinct dimensions of demographic and exposure variation (**Fig. 1C, Fig. S2**). For example, axis 2 correlated with loneliness (*r* = -0.74, p < 1 × 10^−250^), axis 6 with alcohol use and smoking (*r* = 0.87 and 0.32, both p < 1 × 10^−250^, respectively, **Fig. 1C**). While some variables loaded primarily onto single axes (smoking, for instance) others, like spirituality, showed moderate correlations across multiple axes, suggesting their influence spans multiple social and behavioral domains (**Fig. 1C**).

Overall, these results demonstrate that MCA successfully extracts structured, interpretable patterns from sparse and highly correlated social and behavioral determinants of health. The embeddings capture both discrete lifestyle signatures and more diffuse, multidimensional constructs while simultaneously reflecting sociodemographic variation and the non-random survey response patterns typical of biobank studies ^8^. This multifaceted capture of variation makes MCA embeddings particularly suitable for comprehensive disease risk modeling.

### MCA embeddings reveal key contributors to disease risk and help quantify their relative impact on outcome without requiring prior assumptions

Next, we investigated whether MCA embeddings can be used to improve disease risk prediction and how their predictive value compares with commonly used demographic factors and established measures of genetic risk. To this end, we first divided the All of Us cohort into training (80%) and test (20%) sets. We used the training set to train models to predict disease status for each disease using the following predictors: age, gender (excluding gender for prostate and breast cancer models, which affected only men or women, respectively), PGS, 15 population principal components (PCs), and the first 20 MCA embeddings (**Methods**).

Given the genetic diversity of our cohort (**Table 1**) and the known variability in PGS performance across ancestry groups ^9^, we constructed PGS using variants and effect sizes recently reported by the eMERGE network, which were selected and calibrated for optimal cross-ancestry PGS transferability and performance ^4^. As expected, higher scores were associated with increased disease risk (**Fig. S3**). Concordant with results from eMERGE ^4^, calibrated PGS showed similar distributions across genetic ancestry groups but slightly better predictive power in individuals of European genetic ancestry (**Fig. S4**).

To avoid collinearity in our models, we assessed the correlation of each MCA axis with other covariates. The maximum absolute observed correlation was MCA axis 1 correlation with genetic PC1 and PC2 (*r* = 0.30 and 0.32, respectively, both p< 1 × 10^−250^, **Fig. 2A**). Correlations between other ancestry PCs and MCA axes ranged in magnitude from 4 × 10^−5^ to 0.30, indicating that MCA embeddings can be included along with our above-mentioned covariates in our models (**Fig. 1C & 2A**).

**Fig. 2.**
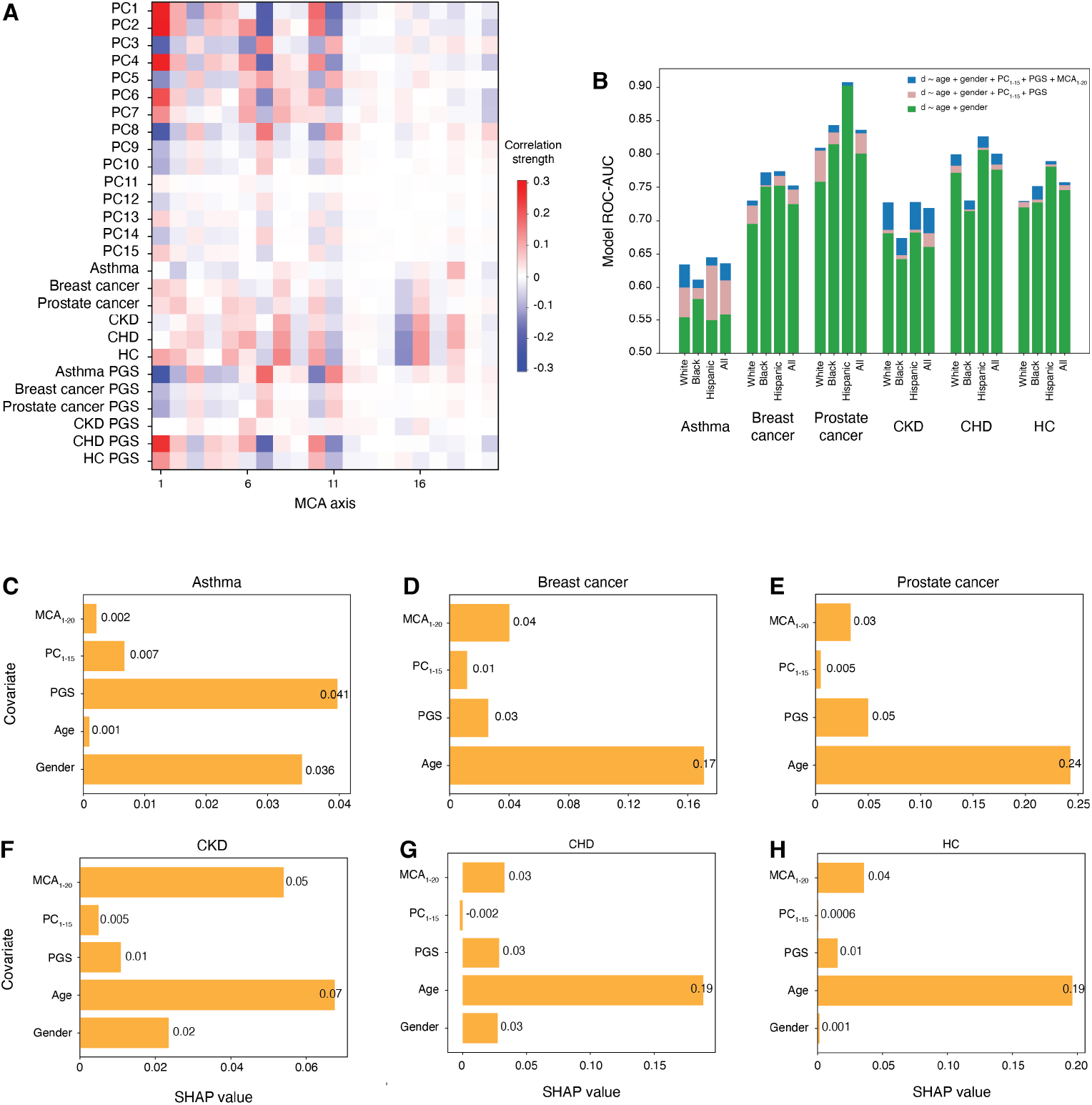
Predictive performance and feature importance across six diseases. **(A)** Heatmap of correlations between the first 20 MCA axes, genetic PCs, disease status, and polygenic scores (PGS). Colors represent correlation magnitude, with Pearson’s correlation used for continuous traits and the point-biserial correlation for binary disease status. **(B)** Predictive power of models, quantified as test-set ROC-AUC in classifying disease (d) status using a specific model. Adding MCA embeddings increased predictive performance across all diseases (ΔAUC = 0.006-0.04) beyond demographics and genetic factors alone, with similar gains observed within self-reported race/ethnicity groups. Y-axis: shows the total AUC for each model colors show which covariates are included in each model. **(C-H)** Relative predictor importance, estimated using Shapley additive explanation (SHAP) values in the test set, for **(C)** asthma, **(D)** breast cancer, **(E)** prostate cancer, **(F)** chronic kidney disease (CKD), **(G)** coronary heart disease (CHD), and **(H)** hypercholesterolemia (HC). SHAP values indicate the average contribution of each feature to model ROC-AUC.

We first evaluated overall model performance in the test set using the area under the receiver operating characteristic curve (ROC-AUC; **Methods**). ROC-AUC values ranged from 0.64 for asthma to 0.84 for prostate cancer (**Fig. 2B, Fig. S5**). These patterns remained robust across self-reported race/ethnicity groups (**Fig. 2B**), which are a better proxy for environmental, behavioral, and social determinants of health than genetic ancestry groups ^10,11^.

We then applied Shapley additive explanations (SHAP) to quantify the relative contribution of each predictor (**Fig. 2C-H**). Age was the strongest predictor for prostate cancer (SHAP = 0.24), hypercholesterolemia (0.20), coronary heart disease (0.19), breast cancer (0.17), and chronic kidney disease (0.07). MCA embeddings were the second most important predictors for chronic kidney disease (0.05), breast cancer (0.04), hypercholesterolemia (0.04), and coronary heart disease (0.03). For asthma and prostate cancer, PGS contributions (0.05 and 0.04, respectively) exceeded those of MCA (0.03 and 0.002, respectively), with PGS being the largest contributor for asthma. Overall, these results are consistent with the age-related nature of the complex disease we studied here ^12–15^, and their higher prevalence among older individuals in our cohort (**Table 1**). They also demonstrate that MCA embeddings provide meaningful improvements across several diseases, highlighting the additive and complementary value of demographic, genetic, and social-behavioral context in disease risk prediction. The relatively low contribution of MCA embeddings to asthma may reflect limitations in the available variables to capture fine-scale known environmental risk factors (e.g., pollutant levels), as well as the overall lower predictive performance of our asthma risk model (ROC-AUC = 0.64) compared with other diseases.

Individual MCA axes showed disease-specific patterns of contribution. While different axes were important for different traits, several consistent patterns emerged (**Fig. 3A**). Axis 18, correlated with the area deprivation index (*r* = -0.37, p < 1 × 10^−250^, **Fig. 1C**), strongly influenced asthma, chronic kidney disease, and coronary heart disease risk (**Fig. 3A**). Axis 8, followed by axes 15, and 16 were primary contributors to hypercholesterolemia as well as chronic kidney disease and coronary heart disease, and risk (**Fig. 3A**). These three axes are correlated with age (*r* > 0.30, p < 1 × 10^−250^, **Fig. 1C**), which may explain part of their effect. Breast and prostate cancers showed strong associations with MCA axis 2 (**Fig. 4A**), which also correlates strongly with loneliness (r = -0.75, p <1 × 10^−250^, **Fig. 1C**). These associations translated into clear clinical relevance: stratifying individuals by deciles of the top contributing MCA embeddings revealed pronounced gradients in disease risk across the full dataset (**Fig. 3B-G**), similar to the well-established increases in prevalence observed across PGS deciles (**Fig. S4**).

**Fig. 3.**
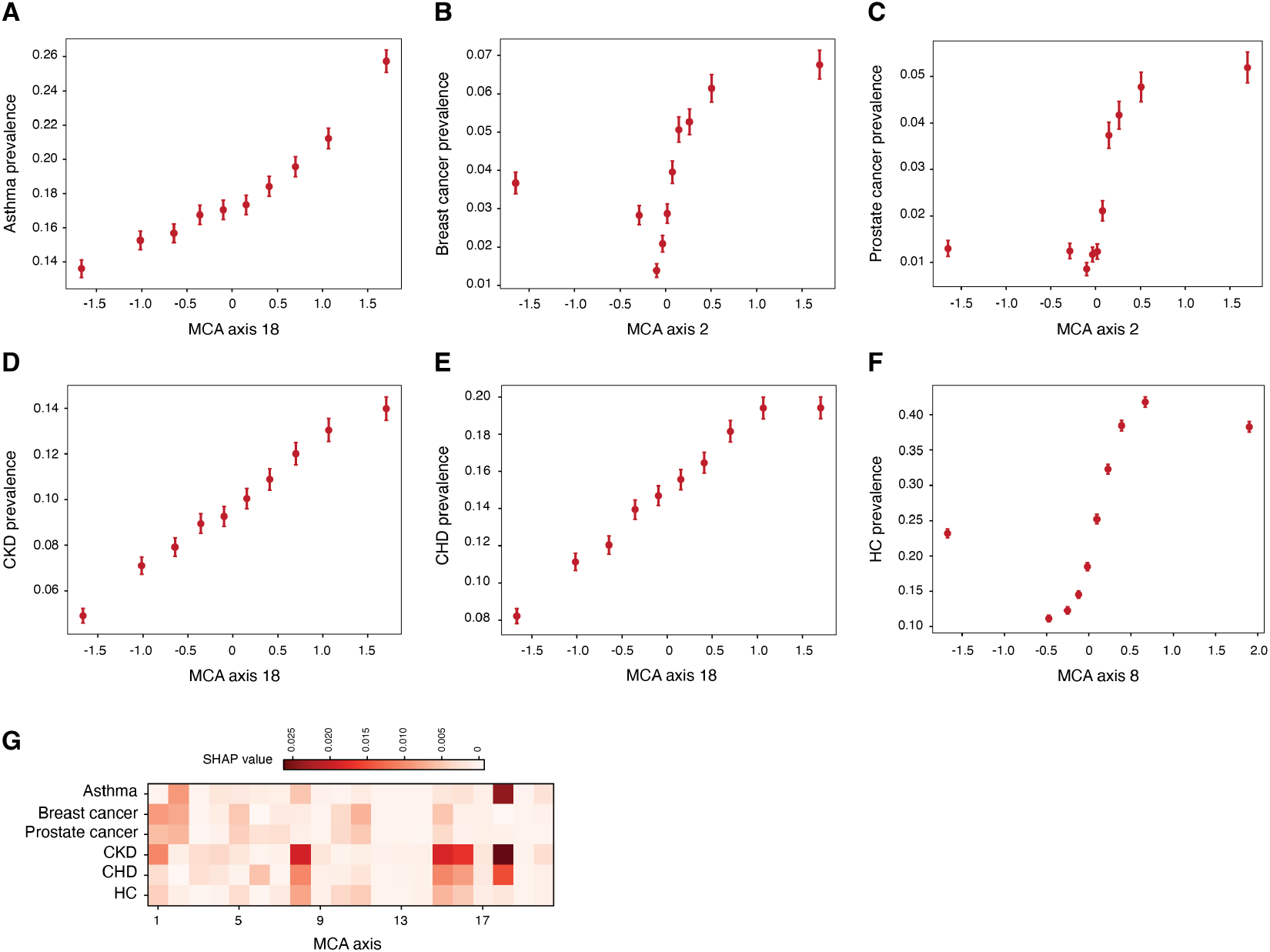
Disease prevalence and MCA axis contributions to predictive performance. **(A-G)** Observed proportion of cases along the MCA axis with the strongest influence on model performance for each disease: **(A)** asthma (axis 18), **(B)** prostate cancer (axis 2), **(C)** breast cancer (axis 2), **(D)** chronic kidney disease (CKD, axis 18), **(E)** coronary heart disease (CHD, axis 18), and **(F)** hypercholesterolemia (HC, axis 8). **(G)** Heatmap of SHAP values showing the relative contribution of each MCA axis across diseases in models including age, sex, PGS, genetic PCs 1-15, and MCA axes 1-20 as covariates.

These findings underscore both the heterogeneous nature of risk factors across complex diseases and the broad utility of MCA embeddings as a tool for improving risk assessment. The embeddings successfully capture meaningful, disease-relevant dimensions of non-genetic risk in a compact form suitable for integration into clinical risk prediction models. The prominent role of age in our models reflects both the age distribution of the All of Us cohort and well-established epidemiological patterns; cancer and cardiovascular disease disproportionately affect older adults, while asthma occurs across the lifespan ^12,13,16,17^. Importantly, the consistent performance across self-reported race/ethnicity groups demonstrates that our model’s effectiveness is not driven by effects that may be specific to socially and culturally constructed population subgroups, supporting its potential for equitable risk assessment.

### Genetic and social/behavioral determinants of disease risk act largely additively, with minimal evidence for interaction

The relationship between genetic predisposition and environmental context has long been hypothesized to interact with environmental, social, and behavioral factors to shape disease risk ^18,19^. To test this possibility systematically in our cohort, we extended our prediction models to include interaction terms between PGS and each of the first 20 MCA embeddings.

Across all six diseases, incorporating these interaction terms produced statistically significant but clinically negligible improvements in predictive performance (**Fig. S5**). The small magnitude of these gains suggests that genetic and non-genetic risk factors, as captured by MCA embeddings, act largely additively rather than synergistically. Furthermore, individual PGS×MCA interactions contributed relatively uniformly across axes, indicating no particular social or behavioral domain disproportionately modifies genetic risk.

We then examined whether accounting for environmental, behavioral, and social context might alter our understanding of the genetic architecture of disease itself. To this end, for each disease, we conducted two parallel genome-wide association studies (GWAS): one using baseline covariates (age, sex, ancestry PCs, genetic relationship matrix, and age-sex interaction terms) and another incorporating these same covariates plus the first 20 MCA embeddings (**Fig. S6-7, Methods**). The variant effect size estimates from these two approaches showed remarkably high concordance for all six diseases (*r* > 0.98, p < 1 × 10^−16^ for all diseases, **Fig. S8-S13**).

Together, these analyses support a predominantly additive model of disease risk. Showing that while environmental, behavioral, and social determinants of health, as captured by MCA embeddings, make substantial independent contributions to disease risk, they do not fundamentally alter genetic risk or variants’ effect sizes.

## Discussion

Complex diseases arise from the joint effect of genetic predisposition and social, behavioral, and environmental context. Leveraging the diversity and depth of the All of Us biobank, we used MCA to convert over 100 categorical environmental, behavioral, and social determinants of health into low-dimensional, interpretable embeddings and asked whether these features (i) improve risk prediction across six common diseases and (ii) modify genetic associations or polygenic risk. MCA embeddings consistently enhanced prediction beyond demographics and PGS, often ranking among the top contributors to model performance. In contrast, adding PGS×MCA interaction terms offered little additional predictive value, and GWAS effect sizes remained highly stable after adjusting for MCA embeddings, indicating limited evidence for G×E at the level of global MCA axes and suggesting that non-genetic context and inherited risk act largely additively in the settings we examined. This additive relationship suggests that interventions targeting social and behavioral factors can reduce disease risk regardless of genetic background, offering hope for broadly applicable public health strategies. These findings also underscore the value of incorporating MCA embeddings in clinical risk models to identify individuals at elevated risk who would be missed by genetic scores or demographics alone.

Methodologically, MCA provides a principled alternative to preselecting a small set of “known” risk factors or relying solely on penalized regression for variable selection. Preselection is hypothesis-driven and risks bias when relevant exposures are unknown ^20^, while regularization tends to retain only the strongest individual predictors without explicitly modeling the relationship between variables, potentially removing variables that are relevant for understanding disease etiology ^21^. By contrast, MCA embeddings summarize correlated categorical inputs into orthogonal, interpretable axes without prior assumptions, and reveal underlying structures that may be missed through assessment of individual covariates ^22–24^. This makes them practical covariates for downstream modeling and sensitivity analyses, conceptually analogous to how genetic principal components summarize population structure.

Our findings align with previous epidemiological studies. Decades of work show that non-genetic factors account for a large share of complex disease burden, often surpassing genetic risk, even for diseases with well-established genetic components, such as asthma, cancers, type 2 diabetes, and neurodegenerative diseases, to name a few ^10,11,19,25^. We likewise observe that MCA embeddings of environmental, behavioral, and social determinants of health contribute substantially to prediction, often more than aggregate polygenic effects. At the same time, we find little evidence for broad gene-environment interaction: PGS×MCA terms minimally affect performance, and variant effect sizes are stable with MCA adjustment. This is consistent with recent reports that MCA-derived social determinants of health have limited impact on heritability estimates ^26^ and that poor PGS portability is unlikely to be driven by pervasive G×E ^27^. These findings do not diminish the importance of environmental, behavioral, or social context for disease risk; rather, they suggest that in these data their influence is largely independent of, rather than modifying, genetic effects.

Several limitations warrant emphasis. Survey nonresponse is incomplete and non-random ^8^, potentially introducing selection bias. Variables included in our MCA analysis were measured retrospectively relative to diagnosis, limiting our ability to make conclusions about how exposures in different stages of life/disease may shape the genetic risk ^25^. Our analyses may be underpowered to detect exposure-specific G×E interactions or effects that require higher-resolution measurements. Additionally, genetic risk modifications that are mediated through higher-level molecular functions, such as epigenetic or transcriptomic processes ^28^ could not be assessed in our framework.

Despite these limitations, our work underscores that while having larger and more diverse genetic cohorts can improve genetic risk prediction, major gains are likely to come from combining genetic data with richer environmental, social, and behavioral context. This can support improved population-level risk stratification and enable more context-aware interventions. Achieving these goals requires closer collaborations between statistical geneticists, genetic epidemiologists, and epidemiologists. Our work also highlights the value of large-scale, diverse, and multimodal biobanks in advancing our understanding of health and disease. Future efforts may focus on harmonizing survey instruments to build transportable low-dimensional summaries of environmental, behavioral, or social determinants of health across cohorts and routinely reporting MCA-style embeddings for these factors alongside genetic PCs. Additionally, incorporating longitudinal data to capture timing and accumulation of exposures and integrating additional data modalities will be essential to fully understand how non-genetic factors shape genetic risk and disease manifestation.

## Methods

### The All of Us biobank

The All of Us Research Program collects detailed health-related data, including demographic information, clinical diagnoses from electronic medical records (EHRs), and genomic data ^29^. We used version C2022Q4R11 of the All of Us database (Controlled Tier Dataset v7 with 413,457 total participants) and filtered for participants with usable whole genome sequence (WGS) data and at least one International Classification of Diseases (ICD) diagnosis code for complex trait cases and controls, leading to 171,614 remaining participants.

### Phenotype selection

We used phecodeX ^30^ to map ICD-9 and ICD-10 codes to phecodes and to identify cases and controls for asthma (phecode RE_475), prostate cancer (CA_107.2), breast cancer (CA_105), chronic kidney diseases (GU_582.2), coronary heart disease (CV_404), and hypercholesterolemia (EM_239.1) using EHRs in the All of Us (N = 171,614). For each phecode, we assigned control status to individuals who did not have a given phecode.

### Survey data

Participants in the All of Us Research Program are invited to complete surveys, including The Basics (25 questions), Lifestyle (26 questions), Health Care Access & Utilization (57 questions), and Social Determinants of Health (80 questions). All survey questions were categorical. For each survey, we performed quality control to select the top 136 questions with the lowest skip and non-response rate, corresponding to a roughly skip/non-response rate of < 30% for post-QC questions. Skip answers included: ‘Skip’. Non-response answers included: ‘Other’, ‘Don’t know’, ‘Invalid’, ‘No matching concept’, ‘None’, ‘Response removed due to invalid value’, and ‘Prefer Not To Answer’. Questions containing information about self-reported sex, gender, race, and ethnicity were excluded from the questions input to the MCA model but were directly included as covariates in some downstream logistic regression models. We also excluded questions about physical disability (e.g., blindness, difficulty walking) because they reflect medical history more than social factors. In addition to survey answers, we also included six different area-level variables based on 3-digit zip codes of participants (i.e., poverty rate, high school education rate, health insurance coverage, assisted income prevalence, median income, and vacant housing prevalence). These area-level quantitative variables were quantized (i.e., turned into categorical variables) before further analysis. We used Cramér’s V to measure the association between pairs of categorical variables

### Multiple Correspondence Analysis (MCA)

We used MCA to explore the linear relationships between our post-QC questions. MCA maps each level of a categorical variable to a separate binary (0 or 1) variable. Because we accept at most one answer from each participant to each question, the resulting indicator matrix contains at most one non-zero entry for all columns associated with one question for one participant. Data was organized in an individual-by-answer matrix of 171,614×575, and MCA was used to summarize and visualize participants’ answers to a set of 50 lower-dimensional composite projections. We used Python 3.10.16, R 4.5.0, and the ade4 package ^31^ (Version 1.7-22) to conduct the MCA analyses using default parameters. We used Point-Biserial correlation to assess the correlation of MCA embeddings with binarized variables and Pearson correlation for quantitative variables such as age and number of questions answered.

### Generating AUDIT-C, ULS-8, and DSES scores

The All of Us surveys we used include the questions from the Alcohol Use Disorders Identification Test (https://www.hepatitis.va.gov/alcohol/treatment/audit-c.asp), UCLA Loneliness Scale (https://labs.dgsom.ucla.edu/hays/files/view/docs/surveys/ULS-8.pdf), and Daily Spiritual Experiences Scale (https://www.dsescale.org/scoring-of-the-daily-spiritual-experience-scale/). For each of these measures, we computed the scores for each participant who had answered all questions using the formula recommended by the corresponding surveys’ creators.

### Polygenic Score (PGS) calculation

We used calibrated variant weights shared by the eMERGE network ^4^ to predict risk for the six conditions. We first lifted eMERGE risk variants from GRCh37 to GRCh38, the reference genome built by the All of Us using Hail (Version 0.2.130). We then calculated individual-level PGS for each condition using code from the AoUPRS library ^32^ using All of Us whole genome sequencing data (N = 171,614). We calculated case prevalence in each PGS decile by dividing the number of cases by the total number of individuals (i.e., cases plus controls) in that decile for each condition.

### Genetic association analysis

#### Genetic data

We used genetic data from All of Us Controlled Tier Dataset v7 to perform genome-wide association studies (GWAS). The v7 release has array data for 312,945 samples using Illumina Global Diversity Array (GDA) and short read whole genome sequencing (srWGS) data for 245,394 samples. All genotype data were mapped to the human genome reference build GRCh38. We assigned individuals to genetic ancestry groups that were pre-calculated for the v7 release using srWGS data. We restricted downstream analyses to African/American (AFR), European (EUR), Hispanic/Latino (AMR), East Asian (EAS), and South Asian (SAS) genetic ancestry groups. Genetic ancestry groups Other (OTH) and Middle Eastern (MID) were removed from every GWAS analysis due to their low case numbers.

#### Principal component analysis (PCA) and genetic relationship matrix (GRM) generation

We used PLINK bed files from array data to obtain PCA vectors and to make a sparse GRM. We performed the following quality control measures on the array data using plink1.9 ^33^: minor allele frequency cutoff of 0.05, individual missingness cutoff of 0.1, Hardy-Weinberg p-value cutoff of 10^-11^, variant missingness cutoff of 0.05, removing individuals with heterozygosity values more than three standard deviations from the sample mean for the ancestry, and removing regions of the genome with long-range high linkage disequilibrium (LD) ^34^. Additionally, for PCA, we LD pruned the array data with an r^2^ threshold of 0.1 using a 50 kb window with a step size of 10 variants. We performed PCA for the European (EUR) group using plink2 –pca approx method ^35^. For other ancestry groups, we perform PCA using the PC-AiR method in the GENESIS R package ^36^, and account for population structure using pairwise relatedness (KING-robust method) calculated using the SNPRelate R package ^37^. To make sparse GRMs, we used SAIGE1.3.6 ^38^, using 5000 random markers, with a relatedness cutoff of 0.125, minor allele frequency cutoff of 0.05, and maximum missing rate of 0.05.

#### Genome-wide association study (GWAS)

We used BGEN files from the Allele Count/Allele Frequency (ACAF) threshold call set for srWGS data as genotype files for performing the association tests. We performed GWAS using SAIGE1.3.6 ^38^ for the following phenotypes if they have at least 20 cases among the genetic ancestry groups that we analyzed (AFR, EUR, AMR, EAS, SAS): asthma, prostate cancer, breast cancer, chronic kidney disease, coronary heart disease, and hypercholesterolemia. Only males were analyzed for prostate cancer, and females for breast cancer. We fit null models (SAIGE Step 1) using the above-mentioned PLINK array data that underwent quality control, using the sparse GRM for the respective ancestry and the following covariates: age, sex, PC1-10, age^2^, sex*age, sex*age^2^ (the covariates with sex were removed for breast cancer and prostate cancer). For GWAS that controls for social determinants of health, 20 MCA embeddings were added to Step 1. We performed single-variant association tests (SAIGE Step 2) using a minor allele count threshold of 200 and with Firth correction for case-control imbalance (p-value cutoff - 0.05). Finally, we performed an inverse variance weighted meta-analysis on genetic ancestry-stratified GWAS results for each phenotype using METAL ^39^.

### Risk prediction

#### Model training

We split the All of Us WGS cohort (N = 171,614) into an 80/20 train/test split. We then used the scikit-learn Python package ^40^ (version 1.3.0) to train an elastic net logistic regression model using three-fold cross-validation while testing different L1/L2 regularization ratios (l1_ratios=[0,.2,.4,.6,.8, 1]) and regularization strengths (Cs=[0.001, 0.01,.1, 1, 10, 100])) to optimize model performance for each of the six common diseases described above. For each disease, the full model included the following covariates: age and gender, the first 15 ancestry principal components (PCs), the PGS value, and the first 20 MCA axes. For breast and prostate cancer, where the analyzed cohort was restricted to women or men, respectively, we did not include gender as a covariate. Regularized logistic regression was used to account for the large number of model covariates, incorporating a penalty term that prevented overfitting to training data and aided extrapolation to unseen test data.

In addition to analysis with our full models, we tested alternate model specifications to validate our chosen features. The alternative models tested are as follows:

1. Condition ∼ age + gender
2. Condition ∼ age + gender + 15 genetic ancestry PCs + PGS
3. Condition ∼ age + gender + 15 genetic ancestry PCs + PGS + 20 MCA embedding
4. Condition ∼ age + gender + 15 genetic ancestry PCs + PGS + 20 MCA embeddings + PGS×MCA embeddings

#### Model testing

We used the test set to compute ROC-AUC for regression models in the overall All of Us cohort and in subsets stratified by self-reported race/ethnicity or genetic ancestry. We compared ROC-AUC values between ablated models that excluded a given set of covariates (e.g., genetics, MCA embeddings, or PGS×MCA embeddings) and models including those covariates, to quantify the incremental contribution of each covariate set to model performance.

#### SHapley Additive exPlanations (SHAP) values

SHapley Additive exPlanations (SHAP) values are widely used in machine learning to explain the output of machine learning models using a game theoretic approach ^41^. Intuitively, SHAP values represent the average marginal contribution of each feature to model performance over all possible model ablations. We used SHAP values generated with our model and the test data to explore the relative importance of different covariates to test set ROC-AUC. The additive property of SHAP values allows us to analyze the impact of individual features (such as a single MCA axis) and groups of features (such as all ancestry PCs).

## Data Availability

The All of Us Research Program oversees researcher compliance with its data management and sharing policies, which are designed to ensure responsible use of data while protecting participant privacy. These policies include tiered data access, the Data and Statistics Dissemination Policy, and the Researcher Code of Conduct. Original data are available only through the All of Us Researcher Workbench to approved investigators. Summary statistics and research results will be disseminated in accordance with journal policies and without restriction at the time of publication.

## Acknowledgements

This work was supported by the following grants: R21MD019104 and R35GM160530. This work was also supported in part through the computational and data resources and staff expertise provided by Scientific Computing and Data at the Icahn School of Medicine at Mount Sinai and supported by the Clinical and Translational Science Awards (CTSA) grant UL1TR004419 from the National Center for Advancing Translational Sciences. Research reported in this publication was also supported by the Office of Research Infrastructure of the National Institutes of Health under award numbers S10OD026880 and S10OD030463. The content is solely the responsibility of the authors and does not necessarily represent the official views of the National Institutes of Health. We gratefully acknowledge All of Us participants for their contributions, without whom this research would not have been possible.

