## Supplementary figures for "A Scalable Framework to Integrate Social Determinants of Health into Disease Risk Models using Biobank Survey Data"

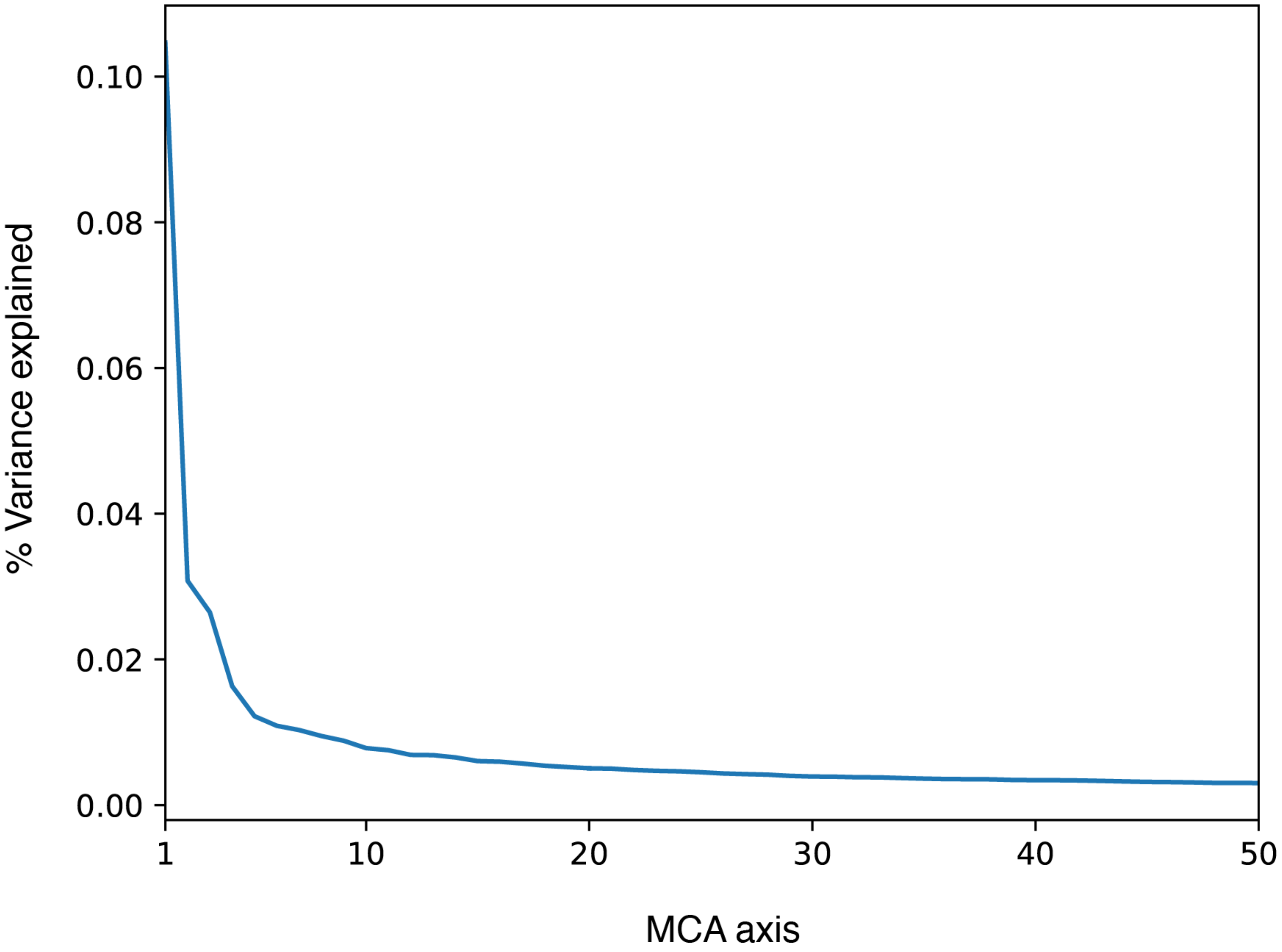


**Fig. S1: Scree plot showing the amount of variance captured by each MCA embedding.** 29.9% of the variance in the input survey data was captured by the first 20 embeddings. Blue color corresponds to the first 20 MCA embeddings and orange to embeddings 21-50.

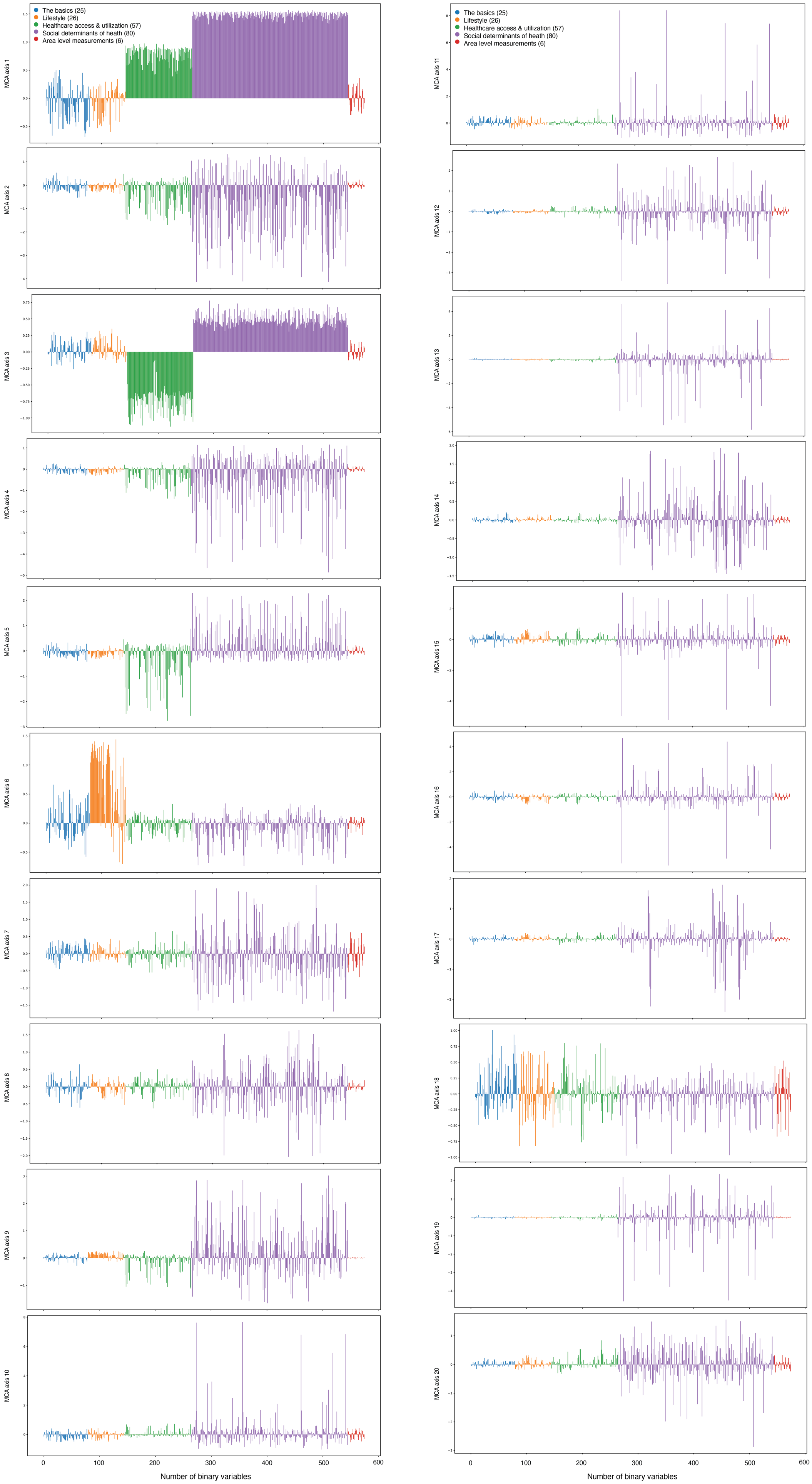

The basics (25)
Lifestyle (26)
Healthcare access & utilization (57)
Social determinants of heath (80)
Area level measurements (6)
MCA axis 1
MCA axis 2
MCA axis 3
MCA axis 4
MCA axis 5
MCA axis 6
MCA axis 7
MCA axis 8
MCA axis 9
MCA axis 10
MCA axis 11
MCA axis 12
MCA axis 13
MCA axis 14
MCA axis 15
MCA axis 16
MCA axis 17
MCA axis 18
MCA axis 19
MCA axis 20
0
100
200
300
400
500
600
Number of binary variables

**Fig. S2: Column projections for MCA embedding 1 to 20.** The X-axis corresponds to binarized survey questions. The Y-axis shows the positions of the survey answer categories in a given MCA axis. Each column projection shows where a particular response (e.g., "yes" to a question, or choosing category *A* vs *B*) lies relative to the other input variables. Categories that are close in the MCA space tend to co-occur across individuals, and categories that are far apart are rarely chosen together.

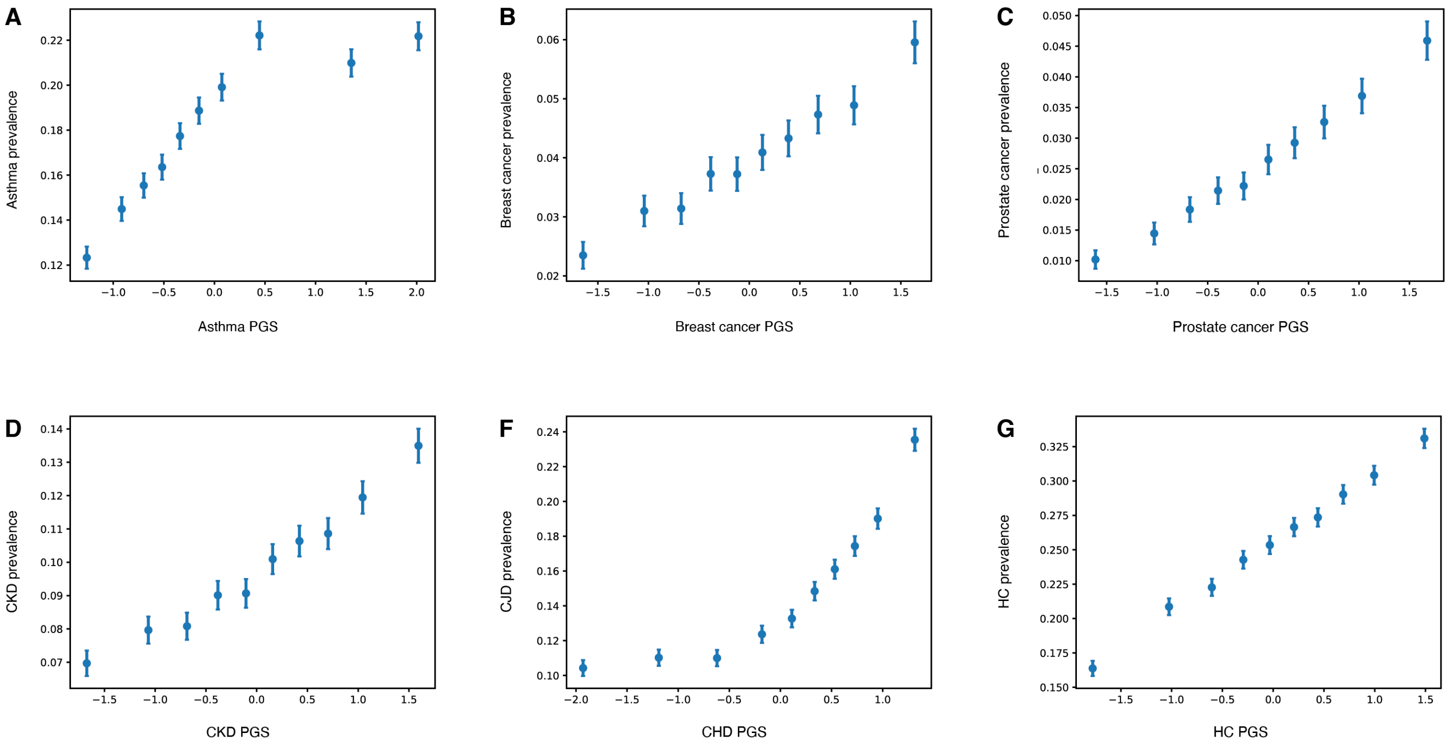


**Fig. S3: Observed proportion of cases across PGS decile. A-F)** Increasing PGS were associated with a higher proportion of cases for (**A**) asthma, (**B**) breast cancer, (**C**) prostate cancer, (**D**) chronic kidney disease (CKD), (**E**) coronary heart disease (CHD), and (**F**) hypercholesterolemia (HC).

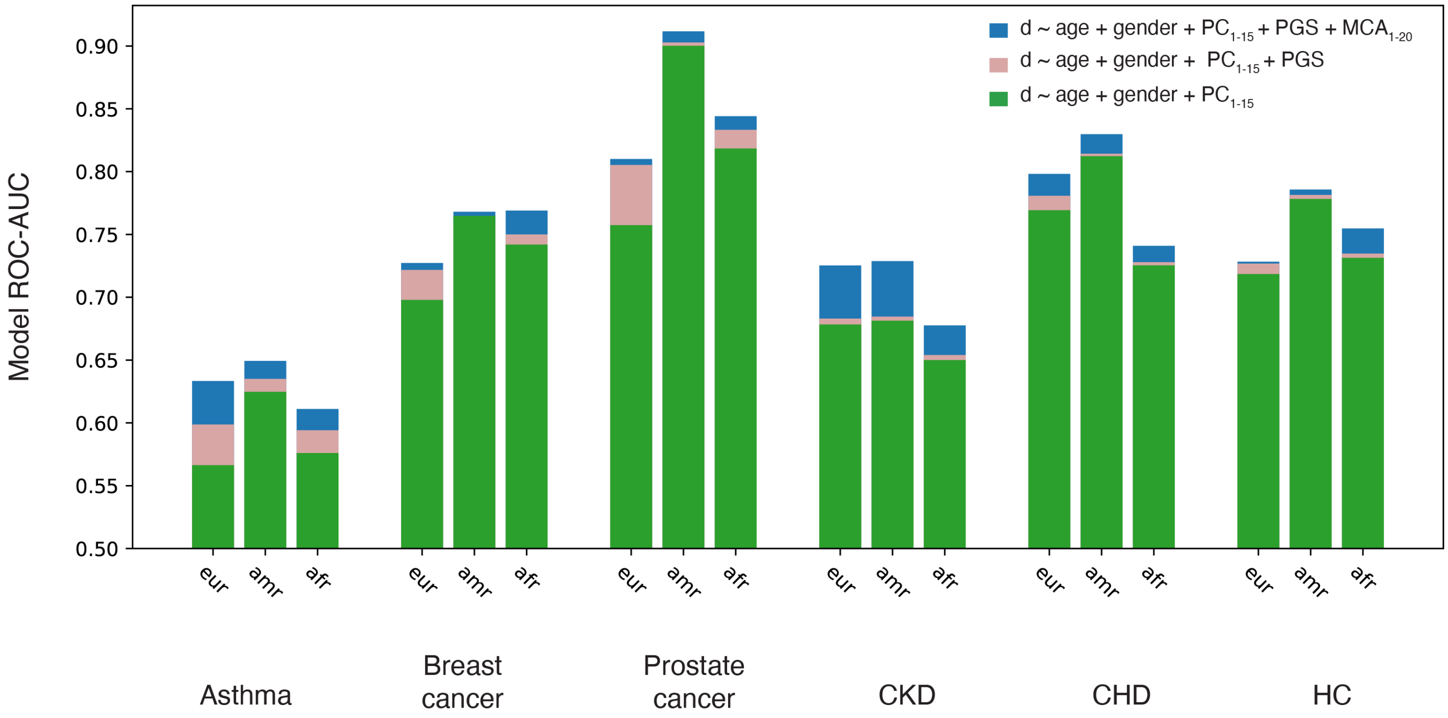


**Fig. S4: Improvement in model performance, assessed using improvement in ROC-AUC in each genetic ancestry group.** Across all traits, including PGS in addition to age, gender, and ancestry PCs 1-15, improved performance with slightly higher improvements in individuals of European genetic ancestry. CKD: chronic kidney disease, CHD: coronary heart disease, HC: hypercholesterolemia, eur: European genetic ancestry, afr: African/African American genetic ancestry, amr: American Admixed/Latino genetic ancestry.

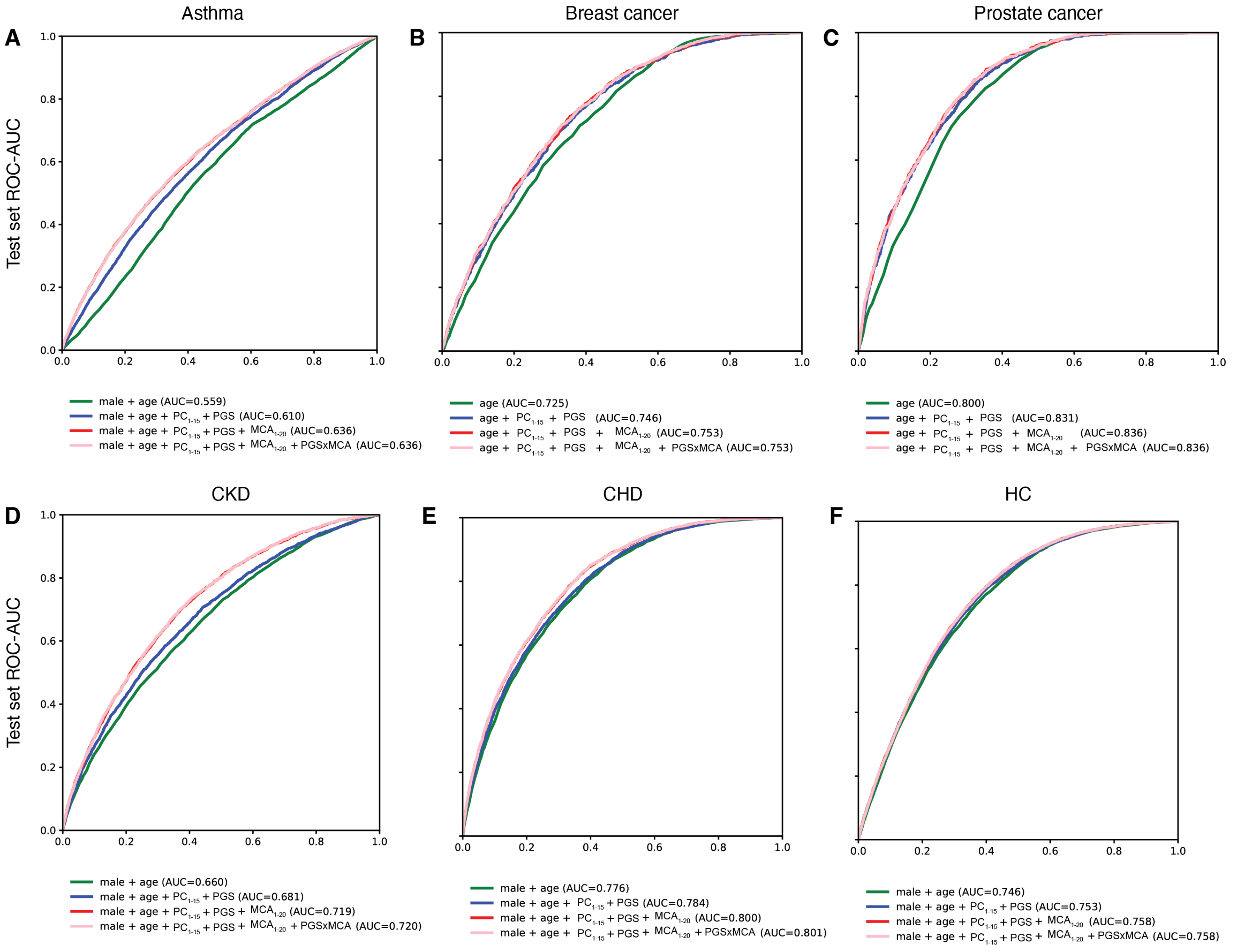


**Fig. S5: ROC-AUC curves for different models for (A)** asthma, **(B)** breast cancer, **(C)** prostate cancer, **(D)** chronic kidney disease (CKD), **(E)** coronary heart disease (CHD), and **(F)** hypercholesterolemia (HC). The overall ROC-AUC ranged from 0.64 (asthma) to 0.84 (prostate cancer). Across all traits, including MCA embeddings, improved model predictive power (measured using ROC-AUC in the test set) beyond what demographic and genetic factors alone could achieve (ΔAUC = 0.006-0.04).

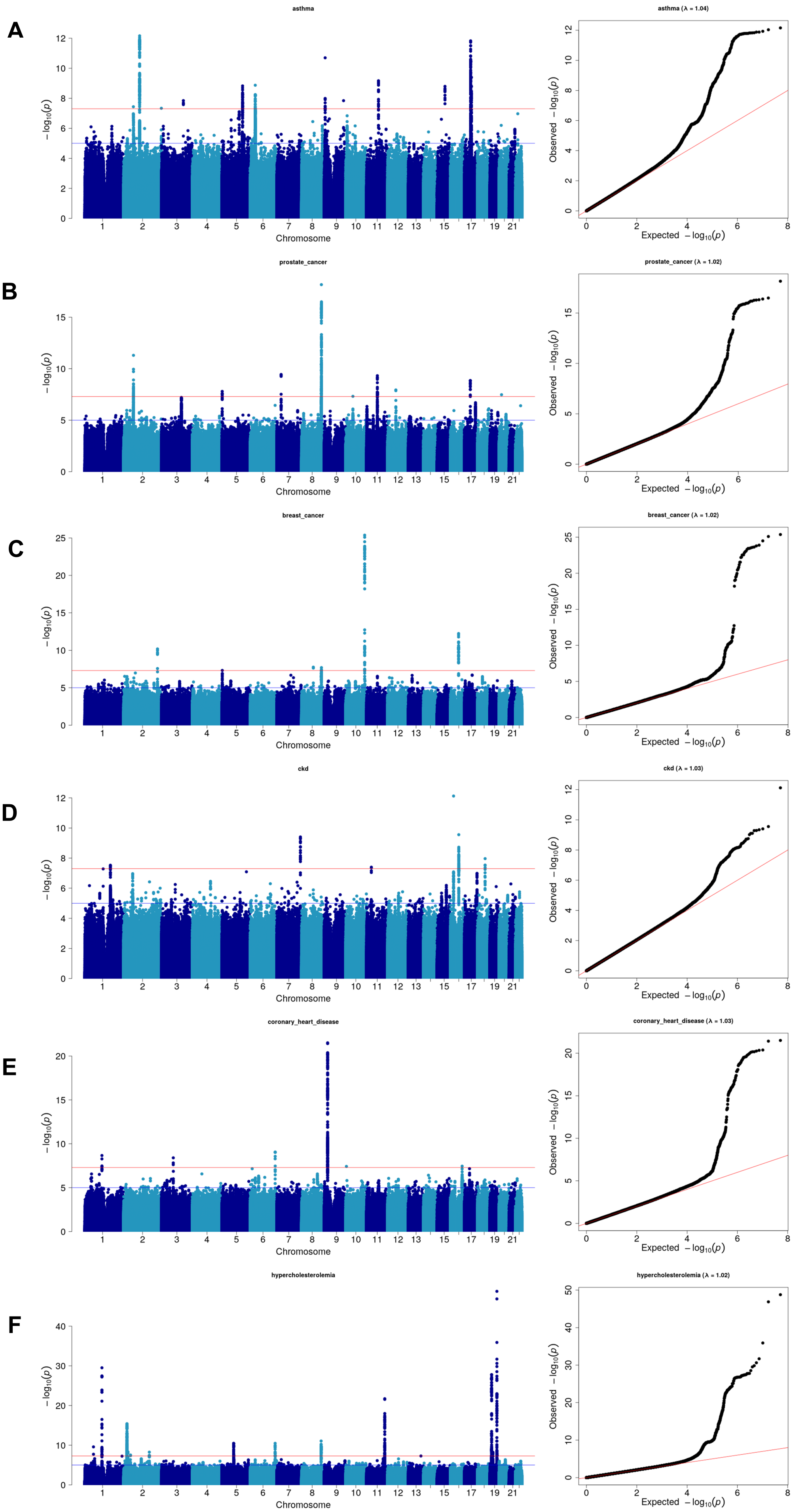
A
asthma
asthma (λ = 1.04)
B
prostate_cancer
prostate_cancer (λ = 1.02)
C
breast_cancer
breast_cancer (λ = 1.02)
D
ckd
ckd (λ = 1.03)
E
coronary_heart_disease
coronary_heart_disease (λ = 1.03)
F
hypercholesterolemia
hypercholesterolemia (λ = 1.02)
$-\log_{10}(p)$
Chromosome
1 2 3 4 5 6 7 8 9 10 11 12 13 15 17 19 21
Observed $-\log_{10}(p)$
Expected $-\log_{10}(p)$

**Fig. S6: Manhattan and QQ plots of genome-wide association study (GWAS) for (A)** asthma, **(B)** prostate cancer, **(C)** breast cancer, **(D)** chronic kidney disease, **(E)** coronary heart disease, and **(F)** hypercholesterolemia**.** GWAS covariates included age, $age^2$, sex, 20 ancestry PCs, age×sex, $age^2$×sex, and a genetic relationship matrix. Ancestry-startified and meta-analysis results are shown. Lambda: genomic inflation factor. X-axis: Chromosome, Y-axis: $-\log_{10}$(association p-value). The vertical line corresponds to the genome-wide significance threshold ($p = 5x10^{-8}$).

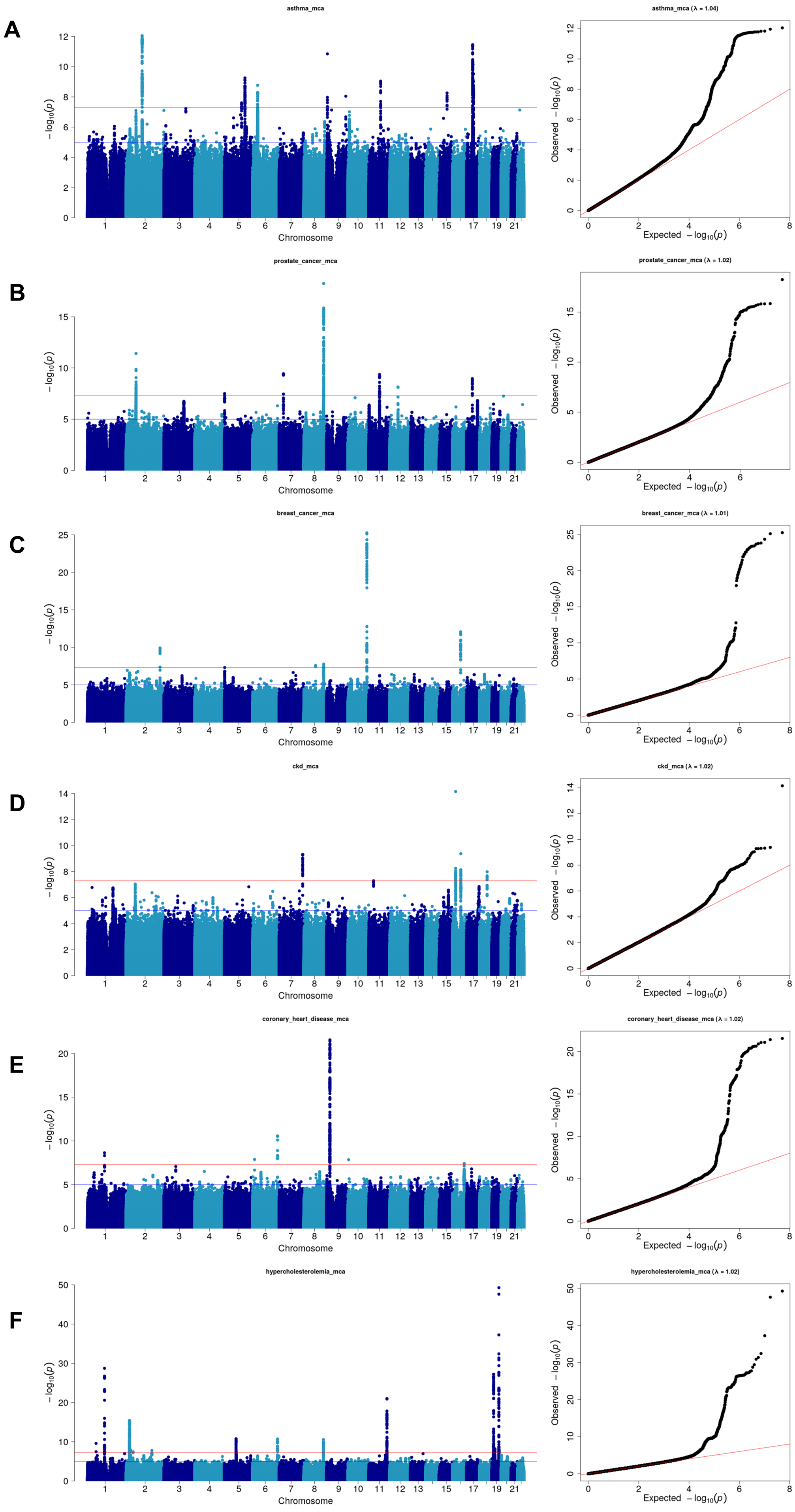
A
asthma_mca
asthma_mca (λ = 1.04)
B
prostate_cancer_mca
prostate_cancer_mca (λ = 1.02)
C
breast_cancer_mca
breast_cancer_mca (λ = 1.01)
D
ckd_mca
ckd_mca (λ = 1.02)
E
coronary_heart_disease_mca
coronary_heart_disease_mca (λ = 1.02)
F
hypercholesterolemia_mca
hypercholesterolemia_mca (λ = 1.02)
$-\log_{10}(p)$
Chromosome
Observed $-\log_{10}(p)$
Expected $-\log_{10}(p)$

**Fig. S7: Manhattan and QQ plots of genome-wide association study (GWAS) for (A)** asthma, **(B)** prostate cancer, **(C)** breast cancer, **(D)** chronic kidney disease, **(E)** coronary heart disease, and **(F)** hypercholesterolemia**.** GWAS covariates included age, $age^2$, sex, 20 ancestry PCs, age×sex, $age^2$×sex, a genetic relationship matrix, and 20 MCA embeddings. Ancestry-startified and meta-analysis results are shown. X-axis: Chromosome, Y-axis: $-\log_{10}$(association p-value). The vertical line corresponds to the genome-wide significance threshold ($p = 5x10^{-8}$).

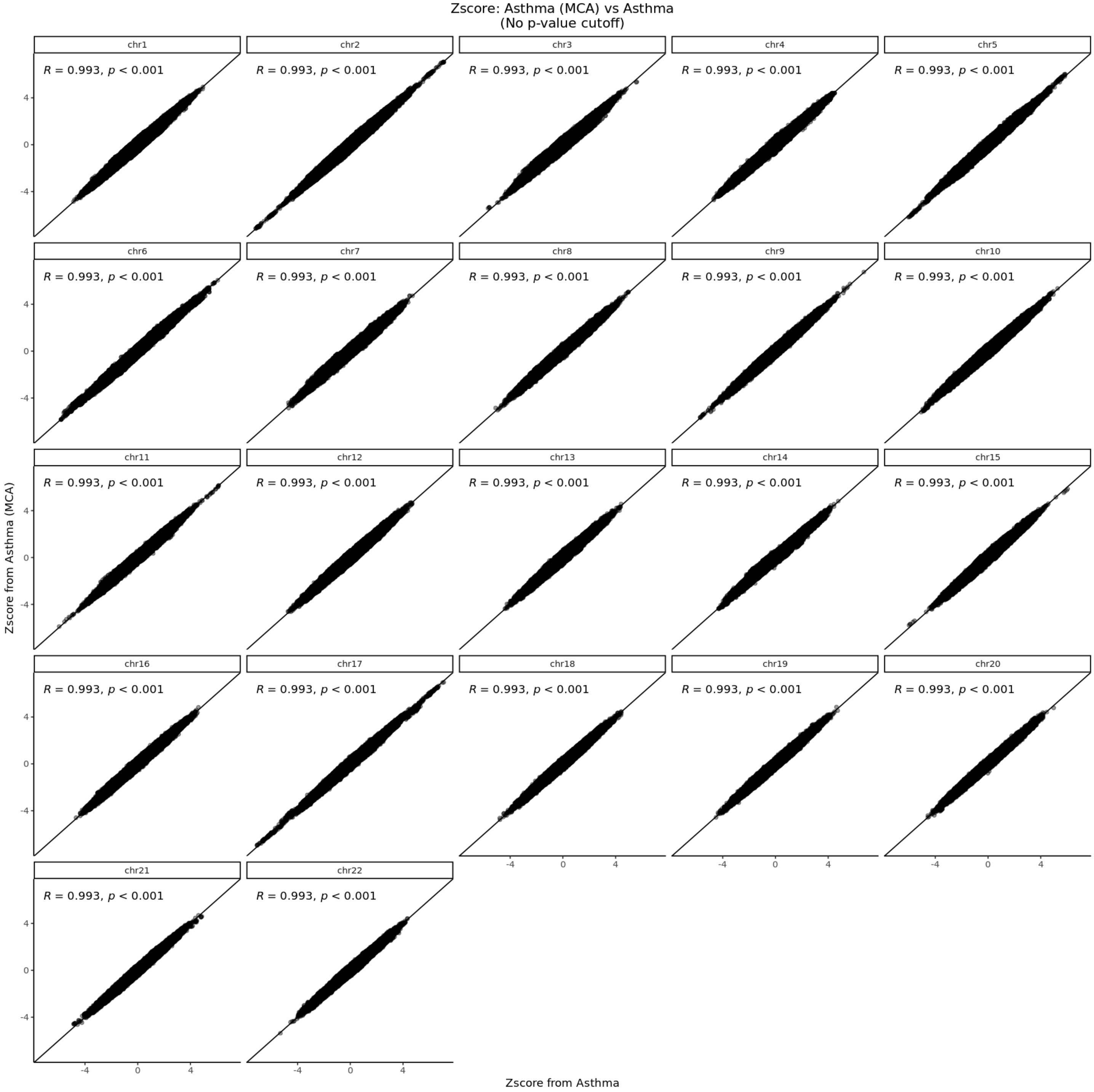


**Fig. S8: Per chromosome effect size correlation for the GWAS with or without MCA embeddings for** asthma. X-axis: effect sizes from the meta-analysis of GWAS that included age, age$^2$, sex, 20 ancestry PCs, age×sex, age$^2$×sex, and a genetic relationship matrix as covariates. Y-axis: effect sizes from the meta-analysis of GWAS that included 20 MCA embeddings in addition to previous covariates. *r* : Pearson correlation coefficient, p: correlation p-value.

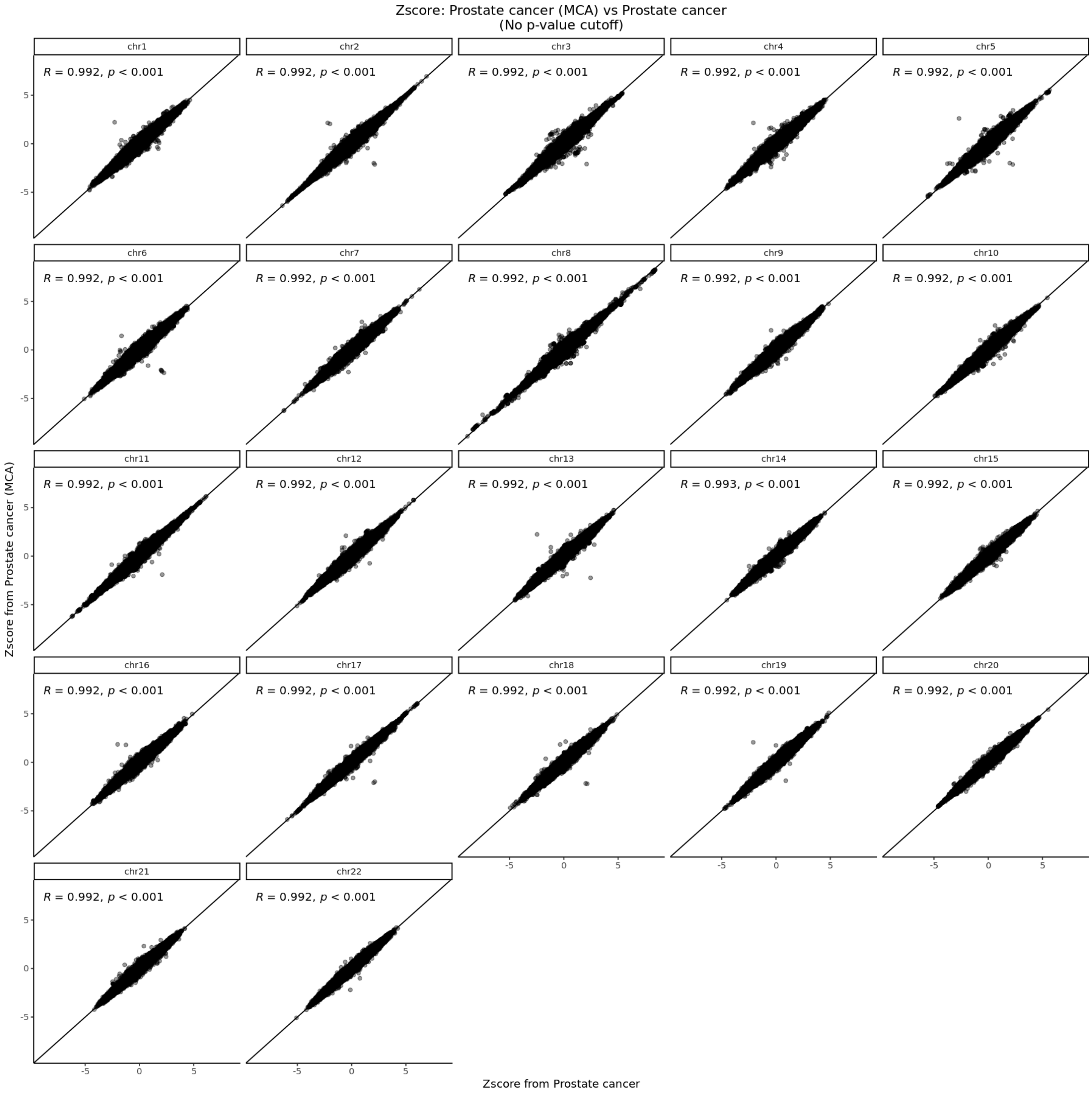


**Fig. S9: Similar to Fig. S8 for prostate cancer.**

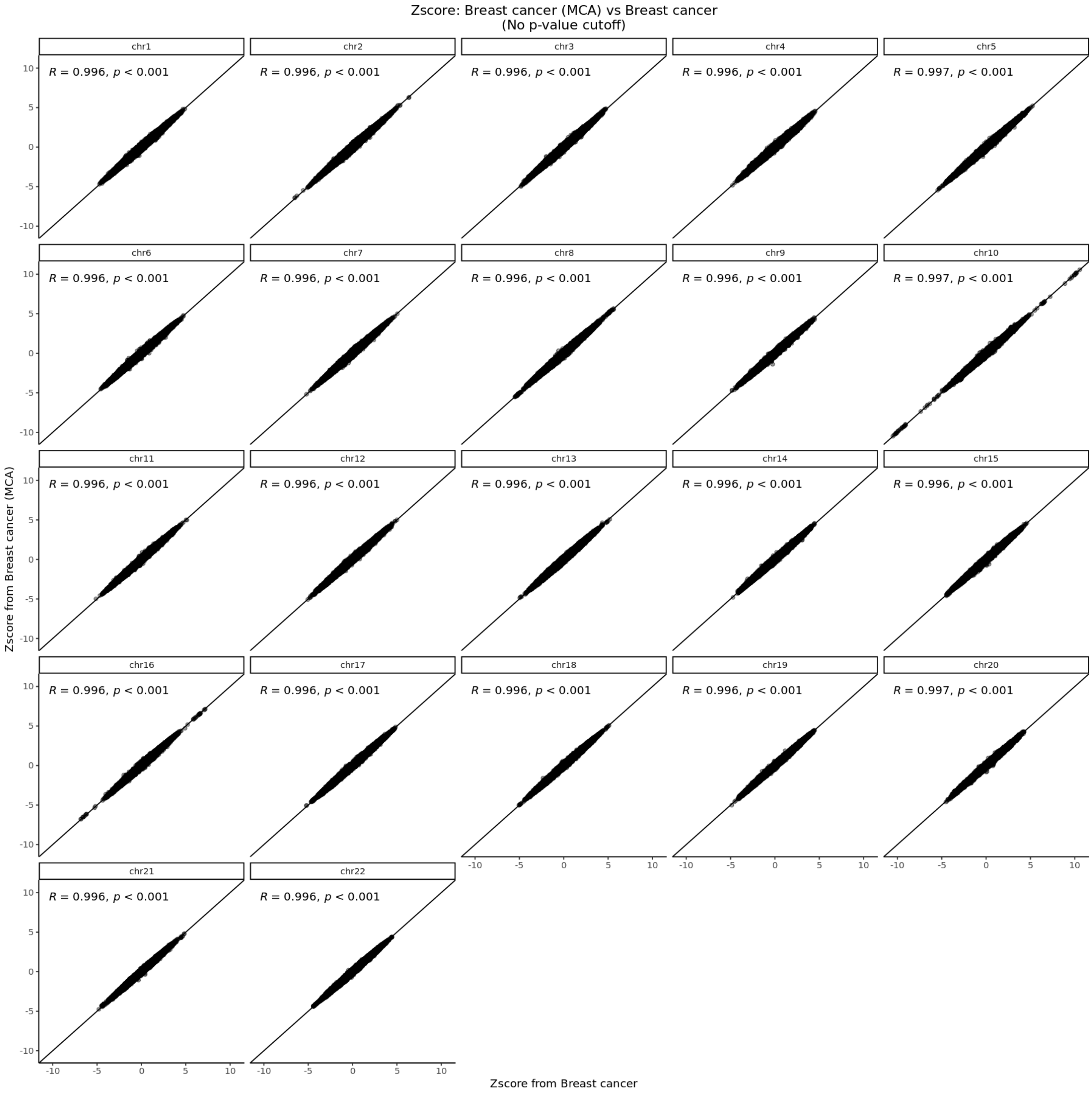


**Fig. S10: Similar to Fig. S8 for breast cancer.**

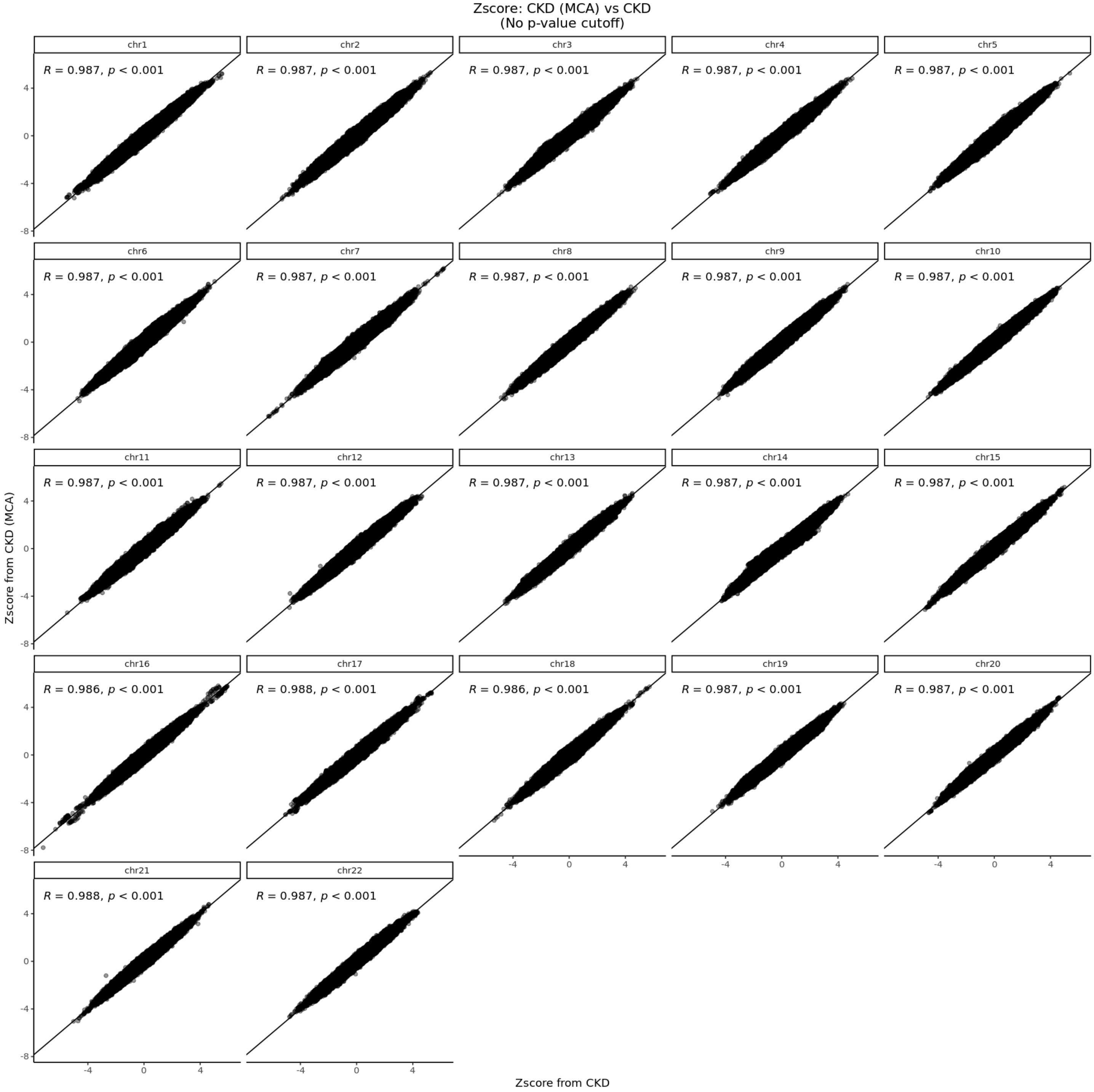


**Fig. S11: Similar to Fig. S8 for chronic kidney disease (CKD).**

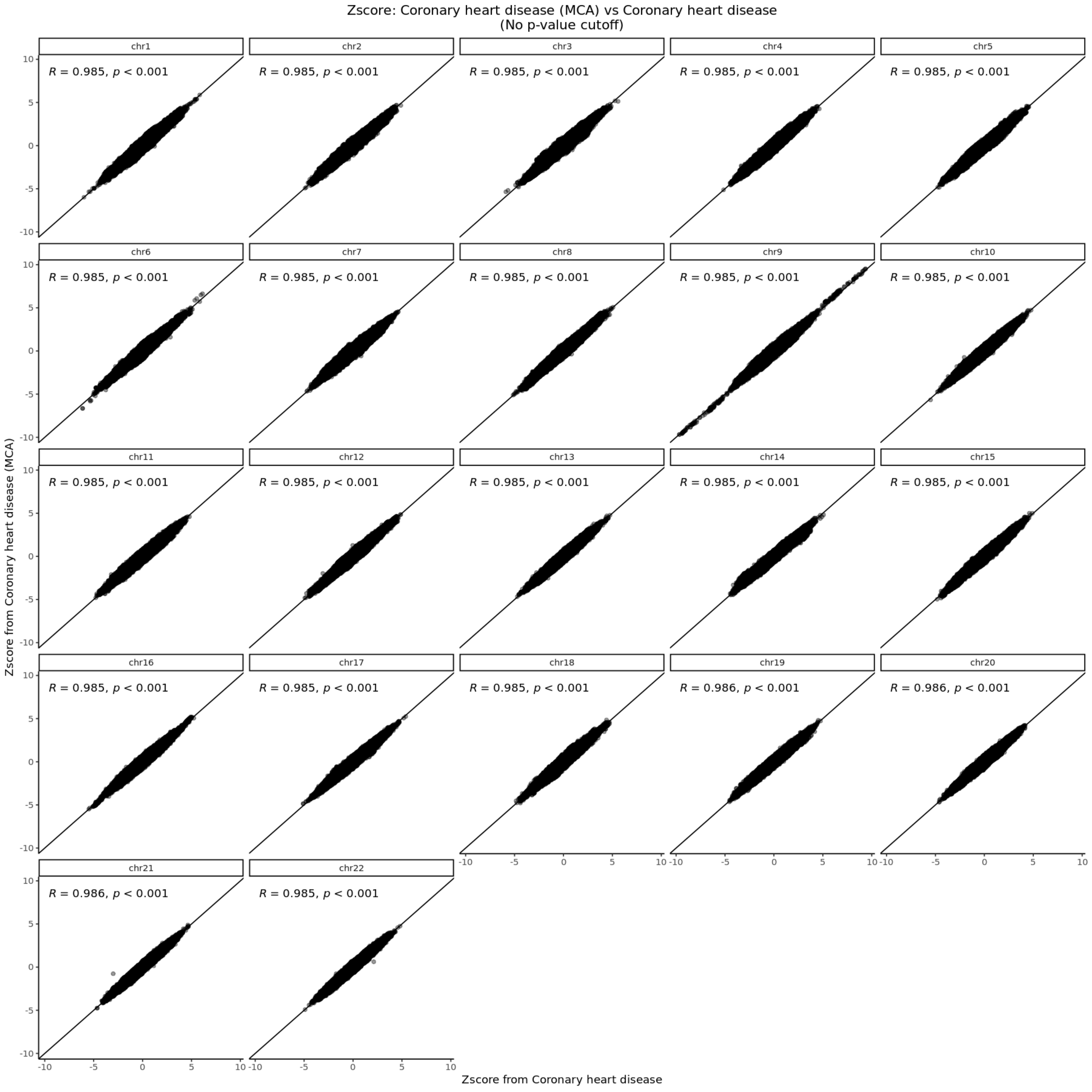


**Fig. S12: Similar to Fig. S8 for coronary heart disease.**

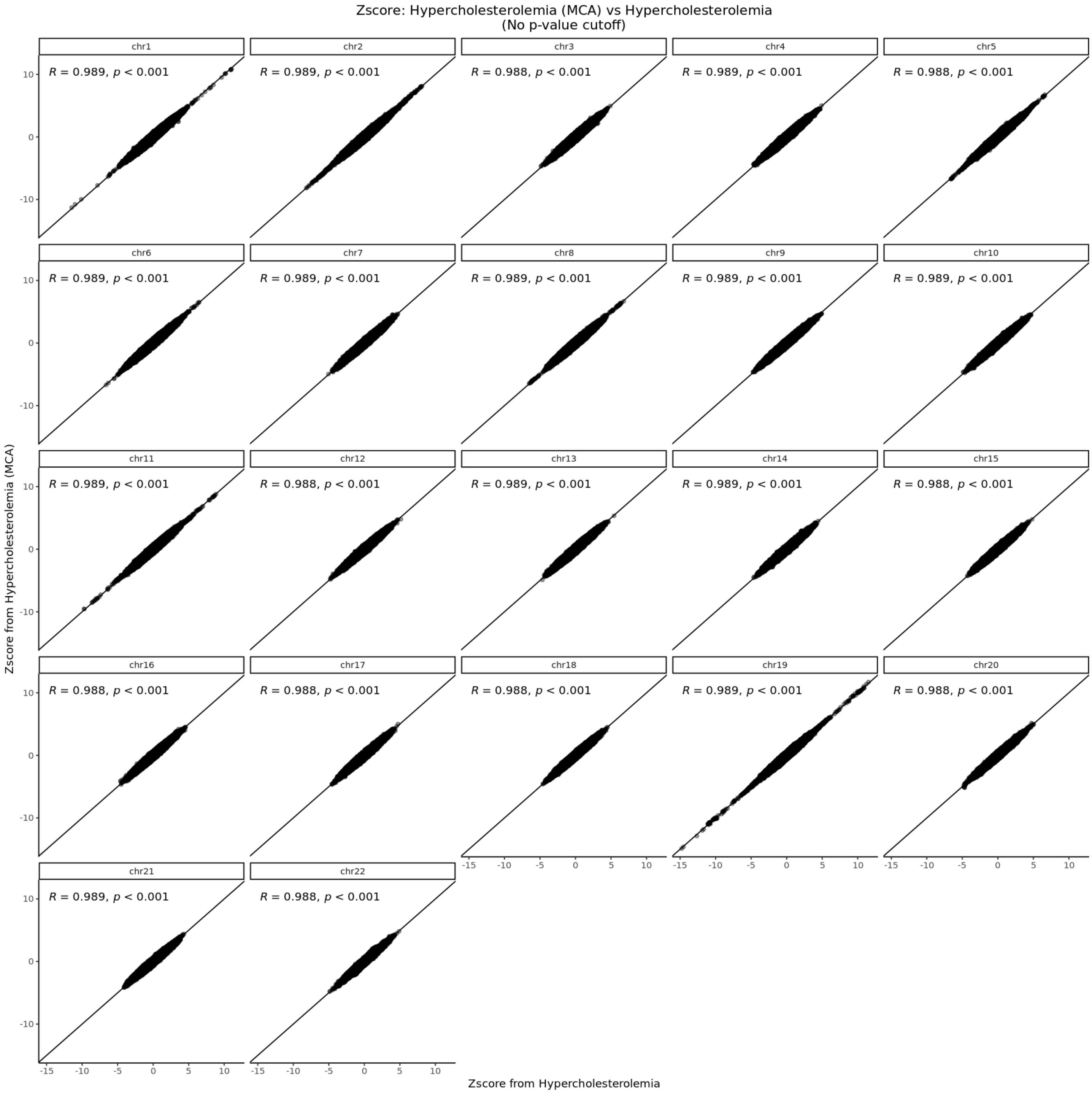


**Fig. S13: Similar to Fig. S8 for hypercholesterolemia**
